# Predicting Motor Trajectories and Mapping Progression Subtypes in Parkinson’s Disease via Structure–Function Neural Field Encoding and Multi-View Representation

**DOI:** 10.64898/2025.12.21.25342791

**Authors:** Shuzhi Zhao, Yushu Zhang, Xu Guorong, Qiao Xue, Yi Pan, Lan Wang, Hanjun Liu, Nan Yan

## Abstract

Parkinson’s disease (PD) is characterized by substantial heterogeneity in progression patterns, posing major challenges for individualized prognosis and clinical management. This study presents Structure–Function Neural Field Alignment and Multi-View Distillation (SFNFA-MVD), a novel deep learning framework that integrates structural, functional, and diffusion MRI to predict motor trajectories and identify progression subtypes of PD. SFNFA-MVD is built around two complementary components: a neural field encoding backbone that maps structural, functional, and diffusion MRI into a continuous structure–function field for fine-grained multimodal alignment, and a multi-view distillation module that leverages transformer-based cross-modal fusion to capture long-range dependencies across modalities and timepoints and generate trajectory-aware clinical predictions. Applied to a longitudinal cohort of 268 PD patients from the PPMI dataset, SFNFA-MVD achieved state-of-the-art accuracy in predicting MDS-UPDRS II and III scores (MAE = 2.167 and 2.164; r = 0.613 and 0.695, p < 0.001), outperforming all benchmarked unimodal and multimodal models. Moreover, unsupervised clustering of fused representations revealed three progression subtypes, slow progressing, moderate progressing, and fast progressing, aligned with distinct alterations in the sensorimotor, dorsal attention, and salience networks, respectively. Importantly, functional network density within each subtype correlated significantly with longitudinal motor decline (r = 0.53–0.57, p < 0.01). These findings demonstrate that SFNFA-MVD enables interpretable, individualized modeling of PD progression, offering a powerful framework for precision prognosis and network-level stratification.

**Author summary:** PD causes gradual loss of movement control and affects people differently, making it hard for doctors to predict how symptoms will change over time. This study developed a new artificial intelligence tool, SFNFA-MVD, that learns from multiple types of brain scans to better forecast individual motor progression. Using structural, functional, and diffusion MRI from 268 people in a long-term Parkinson’s cohort, the model combines information across brain structure and brain activity to predict future motor symptom severity. We found that SFNFA-MVD predicted motor scores more accurately than existing single-scan or simple fusion approaches. Beyond prediction, it also grouped patients into three progression patterns—slow, moderate, and fast—each linked to distinct changes in brain networks involved in movement, attention, and salience processing. Importantly, the degree of network disruption within each group tracked with worsening motor symptoms over time. By connecting imaging-based brain changes with personal symptom trajectories, this approach may help clinicians monitor PD more precisely, identify higher-risk patients earlier, and support more personalized disease management.

## Introduction

Parkinson’s Disease (PD) is a progressive neurodegenerative disorder primarily characterized by motor symptoms such as tremor, rigidity, and bradykinesia [1, 2]. Considering evidence has demonstrated that the trajectory of PD progression varies substantially across individuals, showing significant heterogeneity in the rate and pattern of symptom development [3–5]. Such variability poses challenges for early diagnosis, accurate prognosis, and personalized treatment. Although clinical rating scales such as the Movement Disorder Society Unified Parkinson’s Disease Rating Scale (MDS-UPDRS) [6] remain the primary tools for evaluating symptom severity and disease stage, they rely on subjective ratings and intermittent clinical observation, failing to detect subclinical changes or nonlinear dynamics of disease progression [7–9]. Addressing this gap requires the development of objective, quantitative methods that can accurately detect early brain changes and model individual disease trajectories of PD progression.

A growing body of studies has employed data-driven approaches to characterize model PD progression. For example, probabilistic state modeling has defined discrete motor states and their transitions over time [10], while supervised machine learning has stratified patients into subgroups with distinct rates of motor decline [11]. Other studies have applied methods such as spatial symptom distribution [12] and biomarker-driven clustering [13, 14] to reveal latent heterogeneity in disease trajectories. At the same time, event-based modeling has been used to reconstruct individualized sequences of clinical and neurodegenerative changes [12]. Nevertheless, most models rely primarily on clinical or demographic features and incorporate limited biological data, particularly neuroimaging features that could offer direct insights into brain pathology. They also often assume fixed or linear assumptions on disease trajectories [3, 13], overlooking the non-linear and multidimensional nature of PD progression.

Recent advances have integrated structural and functional magnetic resonance imaging (MRI) with deep learning models to quantify PD severity and predict disease progression [14, 15]. For example, a contrastive variational autoencoder applied to structural MRI (sMRI) identified subcortical and temporal regions associated with MDS-UPDRS scores and longitudinal progression [16]. A graph-based multimodal model integrating sMRI with clinical and genetic data achieved area under the curve (AUC) values of 0.748 and 0.714 for classifying progression status based on Hoehn & Yahr (H-Y) stage and UPDRS-III scores over 12 and 36 months, respectively [13]. Multimodal approaches incorporating T2-weighted MRI and DAT-SPECT achieved an AUC of up to 0.93 for motor progression classification [17] and a classification accuracy of 78-79% in longitudinal progression clustering [18]. Additionally, deep temporal modeling using hybrid CNN-LSTM architectures yielded high accuracy in classifying H-Y stage transitions (AUC ≈ 0.92) [19], while Kolmogorov-Arnold networks outperformed LSTMs in capturing UPDRS trajectories [20]. However, most predictive frameworks rely on simplistic data-fusion strategies, such as simple feature concatenation or late-fusion pipelines [21, 22], that failed to capture the hierarchical interactions across structural, functional, and diffusion modalities. Moreover, while temporal architectures such as CNN-LSTM frameworks have shown promise in predicting short-term stage transitions [23–25], few have applied longitudinal neuroimaging to characterize the individualized nature of PD progression. As a result, current models remain largely constrained to group-level classification or subtype identification [11, 13] rather than individualized prognostic modeling within a multimodal framework.

To address these limitations, we developed a novel deep learning framework, Structure–Function Neural Field Alignment and Multi-View Distillation (SFNFA-MVD, see Figure 1), for individualized modeling of PD progression through multimodal MRI integration. SFNFA-MVD is built around two key ideas: (i) a structure–function neural field encoder that maps sMRI, DTI and fMRI onto a shared cortical manifold to enforce spatially aligned structure–function representations, and (ii) a multi-view distillation module that distills trajectory-relevant patterns across imaging views and timepoints into a unified latent space for individualized PD trajectory prediction, implemented with a local cross-modal encoder and a transformer-based fusion block inspired by recent work [26, 27]. We applied SFNFA-MVD to multimodal MRI data from 268 patients with PD at baseline, 3-year, and 5-year follow-ups in the Parkinson’s Progression Markers Initiative (PPMI). The model was trained to predict longitudinal motor symptoms from MDS-UPDRS Parts II and III and to identify progression subtypes characterized by distinct network-level patterns. We hypothesized that this multimodal MRI fusion framework would achieve superior accuracy in predicting individual motor trajectories and identify clinically interpretable progression subtypes. Proficient in English, consider using an English language editing service before submitting your article. An expert editing service can help you refine the use of English in your article, so you can communicate your work more effectively.

**Figure 1.**
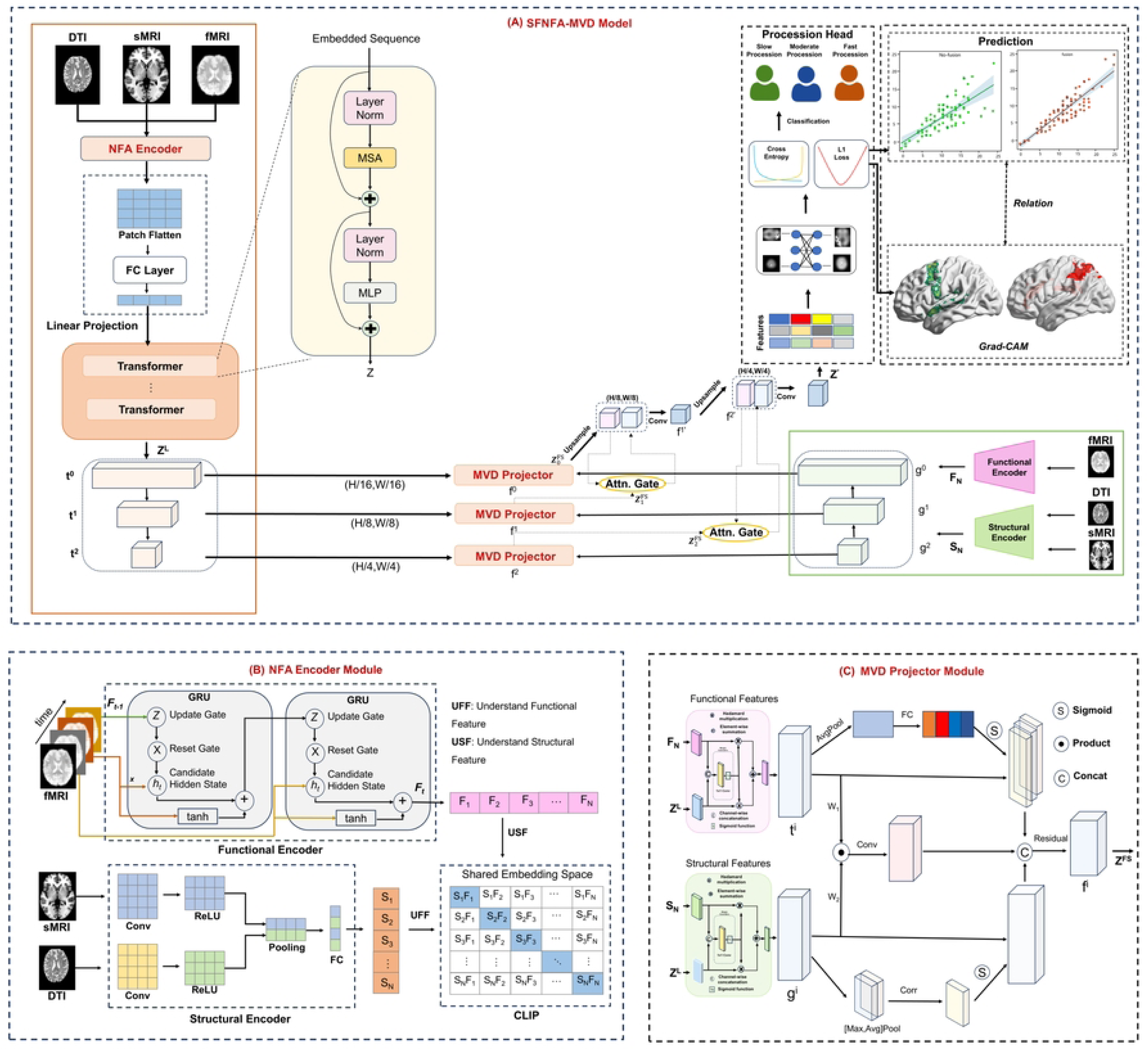
Overview of the SFNFA-MVD framework. *(A)* The SFNFA-MVD framework integrates multimodal neuroimaging inputs (DTI, sMRI, fMRI) through a Dual Encoder and Transformer-based fusion backbone. Each modality is first linearly projected and embedded into a shared latent space, forming hierarchical feature representations 𝑍^𝑙^across multiple scales. Cross-level BiFusion modules and attention gates enable adaptive feature interaction between structural and functional embeddings. The fused representations are decoded for prediction tasks (e.g., regression and classification) via a unified Processing Head, with interpretability provided by relation mapping and Grad-CAM visualization on cortical surfaces. *(B)* The Dual Encoder Module comprises a functional encoder (fMRI) and structural encoder (sMRI/DTI). Each encoder employs convolutional, pooling, and GRU layers to extract modality-specific representations, Understanding Functional Features (UFF) and Understanding Structural Features (USF), which are aligned within a shared embedding space using CLIP-style contrastive learning. *(C)* The BiFusion Module performs bidirectional integration of functional and structural features via attention-guided correlation, concatenation, and residual pathways, producing joint representations 𝑍*_i_^B^* that enhance both global context and fine-grained alignment across modalities.

## Methods

### Experiment Design

#### A. PPMI Dataset

Our model utilized data from the Parkinson’s Progression Markers Initiative (PPMI) dataset [28, 29], a large-scale public resource with 1,400 participants across 11 countries and 33 clinical sites. This study aims to identify motor symptom biomarkers in PD through multimodal imaging. We used multimodal data (N=400) from the PPMI dataset (https://www.ppmi-info.org/), specifically MRI and DTI acquired via the Siemens MAGNETOM Trio 3.0 T MRI scanner. MRI parameters included TR = 2300 ms, field strength = 3 tesla, flip angle = 9°, TE = 2.98 ms, slice thickness = 1 mm, pulse sequence = GR/IR, and acquisition plane = sagittal. DTI parameters were TR = 600– 1000 ms, field strength = 3 tesla, flip angle = 90°, gradient directions = 64, TE = 88 ms, slice thickness = 2 mm, and pulse sequence = EP. Among these data, 268 PD patients [11] provided multimodal MRI data at 0, 3, and 5 years.

#### B. Setup

The SFNFA-MVD model was implemented using the PyTorch toolkit and employed a 5-fold cross-validation strategy (https://github.com/zhaoshuhzi/PD_PPMI_S-F_Fusion) to ensure the model’s robustness and reliability. Model parameters were optimized using the Adam optimizer, with gradient descent and L1 loss functions to enhance learning. The network configuration included a learning rate of 10^-5^, a dropout rate of 0.35, and a batch size of 2. Model performance was evaluated using mean absolute error (MAE), along with CS2 (probability of MAE < 2) and CS3 (probability of MAE < 3) metrics. Additionally, we evaluated various single-modal and multi-modal feature models, including RegNet [30], GRU [31], VGG [32], ResNet [33], DenseNet [34], CLIP[26], and TransFuse [27]. To enhance the interpretability of the model in medical applications, we used Gradient-weighted Class Activation Mapping (GradCAM) [35] to visualize heatmaps overlaid on the original images, highlighting regions that significantly influenced the model’s predictions. This method helps improve the model’s transparency, making it easier for clinicians to understand the rationale behind the model’s decisions.

### Overview of the SFNFA-MVD Framework

SFNFA-MVD (Structure–Function Neural Field Alignment and Multi-View Distillation, see Fig. 1) is a multimodal brain imaging fusion framework designed for structure–function consistency modeling, aiming to unify the extraction, alignment, and integration of complementary information from sMRI, DTI, and fMRI. Its core is a structure–function neural field encoder that first learns a structural neural field basis from sMRI and DTI and then uses it to guide the dynamic encoding of fMRI. Convolutional encoders on structural modalities discover the most informative anatomical bases, while a GRU-based functional encoder leverages these bases as priors to predict future functional activity from past dynamics, explicitly enforcing structure–function consistency. To verify and regularize the stability of this structure-to-function encoding across subjects, SFNFA-MVD adopts a CLIP-style contrastive objective that not only aligns sMRI and fMRI, but also constructs a shared structure–function neural field space. On top of this encoder, a Transformer is embedded into a U-Net backbone to jointly model local and global dependencies in this shared space, yielding multi-scale, globally-aware representations. Finally, a multi-view distillation with BiFusion blocks projects the unified representation at multiple scales from two complementary views, enhancing subtype-sensitive spatiotemporal complementarity. Overall, SFNFA-MVD is organized around structure–function consistency learning, with coordinated encoding, Multi-View Distillation, and attention mechanisms to improve disease progression modeling and key region identification.

#### A. Structure–Function Neural Field Encoder and Shared Latent Space Mapping

To align multimodal images, SFNFA-MVD introduces a structure–function neural field encoder that extends the classic dual-encoder idea. The encoder extracts features from structural (sMRI, DTI) and functional (fMRI) modalities and maps them into a shared structure–function semantic space using contrastive learning.

On the structural side, CNN-based encoders learn a structural neural field basis, namely a compact set of anatomical feature bases capturing dominant patterns of cortical atrophy, subcortical changes, and white-matter alterations. On the functional side, a GRU-based encoder models temporal dependencies of fMRI conditioned on this structural basis.

Intuitively, the structural neural field provides a coordinate system in which past functional activity can be used to predict subsequent activity, enforcing structure–function consistency in the learned dynamics.

##### 1.1 Functional Encoder

fMRI reflects temporal brain dynamics, requiring models that capture sequential dependencies. We use GRU as the functional encoder to effectively extract temporal features. Its reset and update gates balance memory and new input, supporting PD progression modeling. At time step t, the reset gate is computed as:

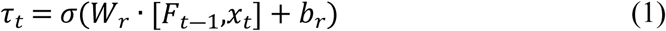

where 𝜎 is the sigmoid activation function, 𝑊_𝑟_ is the weight matrix of the reset gate, 𝐹_𝑡―1_ is the previous hidden state, 𝑥_𝑡_ is the current input, and 𝑏_𝑟_is the bias of the reset gate. The update gate controls how much past and new information is kept, ensuring smooth flow and stable, efficient training. It is computed as:

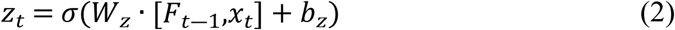

where 𝑊_𝑧_ and 𝑏_𝑧_ are the weight matrix and bias term of the update gate, respectively.

The candidate hidden state combines the current input and reset-modulated past state, forming the basis for the final hidden state after update gate adjustment. It is computed as:

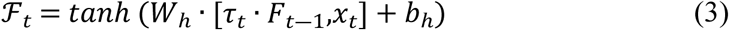

where 𝑊_ℎ_ and 𝑏_ℎ_ are the weight matrix and bias term of the candidate hidden state, respectively. The hidden state is then updated as follows:

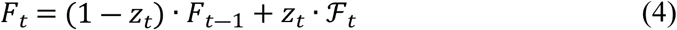

The update gate 𝑧_𝑡_ controls the fusion of the previous hidden state 𝐹_𝑡―1_ with the new candidate hidden state ℱ_𝑡_.

##### 1.2 Structural Encoder

To extract high-resolution anatomical features from sMRI and DTI, we employed a CNN-based encoder. CNNs effectively capture spatial patterns such as cortical atrophy, subcortical changes, and white matter alterations—key to understanding PD-related structural changes. The encoder includes stacked convolutional layers with activations, pooling, and fully connected layers, progressively abstracting intensity into morphological representations. sMRI and DTI are processed through parallel CNN pipelines to learn modality-specific features, which are later fused with functional features. The convolution operation is defined as:

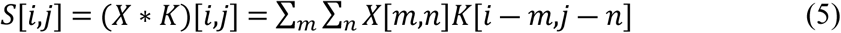

where *X* is the input image, *K* is the convolution kernel, and S[*i*,*j*] is the value at a specific position in the output feature map. To model complex anatomical variations, ReLU activation is applied to introduce non-linearity:

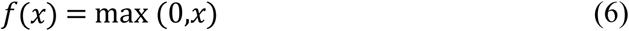

To reduce the spatial dimensions of the feature map, retain key features, and improve noise resistance, pooling operations are applied as follows:

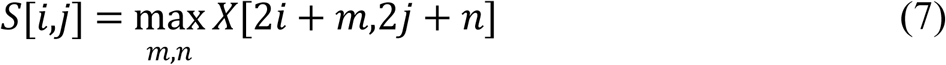

Finally, a fully connected layer extracts the structural features by:

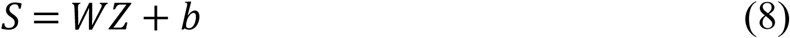

where *Z* is the input feature, *W* is the weight matrix, and *b* is the bias term.

##### 1.3 CLIP-Based Structure–Function Neural Field Alignment

To align structural and functional MRI, we adopt a contrastive learning strategy based on the CLIP framework. CLIP uses a dual-encoder: a CNN for spatial features from sMRI and DTI, and a GRU for temporal features from fMRI. This enables mutual understanding between modalities by mapping them into a shared semantic space. The objective is to maximize similarity between matched pairs and minimize it for unmatched ones, measured by cosine similarity:

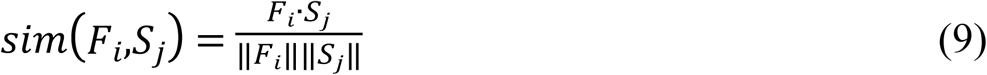

where 𝐹_𝑖_ is the structural image feature and 𝑆_𝑗_ is the functional image feature. To maximize the similarity of structural-functional pairs, the InfoNCE contrastive loss function is introduced as:

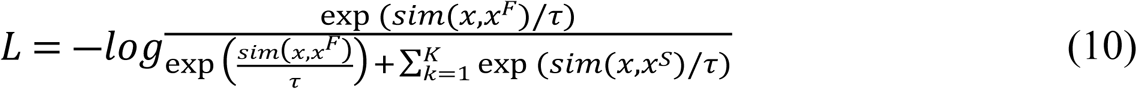

where 𝑥^𝑆^ is the structural image feature, 𝑥^𝐹^ is the functional image feature, and 𝜏 is a temperature parameter that adjusts the similarity distribution range.

##### 1.4 Shared Embedding Space for Structure–Function Matching

Using CLIP’s dual-encoder framework, structural (sMRI, DTI) and functional (fMRI) features are projected into a shared embedding space. This space captures cross-modal similarities, forming a structure–function alignment matrix. The similarity between embeddings is computed as:

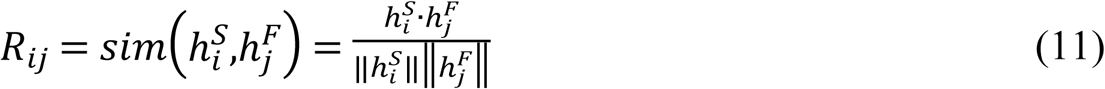

Where 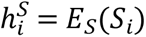 is the embedding representation of structural sample 𝑆_𝑖_, and 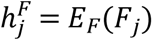 is the embedding of functional sample 𝐹_𝑗_. Here, 𝑆 = {𝑆_1_,𝑆_2_,…,𝑆_𝑛_} represents the structural dataset with 𝑆_𝑖_ as individual samples, and 𝐹 = {𝐹_1_,𝐹_2_,…,𝐹_𝑛_} represents the functional dataset. 𝐸_𝑆_(𝑆_𝑖_) is the structural encoder output for 𝑆_𝑖_, while 𝐸_𝐹_(𝐹_𝑖_) is the functional encoder output for 𝐹_𝑖_.

#### B. Transformer-Based Unified Cross-Modal Representation

To capture long-range dependencies and build unified representations, SFNFA-MVD introduces a Transformer module after the encoders to jointly model structural and functional features.Each Transformer layer includes multi-head self-attention (MSA) and a feed-forward network (MLP) to capture global cross-modal feature relations.

##### 2.1 Self-Attention for Global Alignment

Aligned features from the Dual Encoder are split into patches, flattened, and linearly projected via an FC layer to form Transformer input tokens. Self-attention enables the Transformer to capture long-range dependencies by weighting image patches. Its computation is:

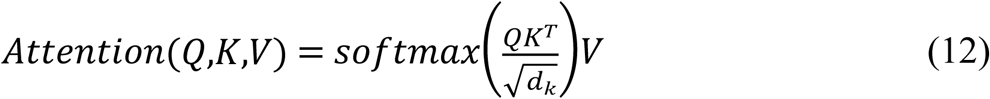

Where 𝑄 = 𝑋𝑊^𝑄^, 𝐾 = 𝑋𝑊^𝐾^, and 𝑉 = 𝑋𝑊^𝑉^are the query, key, and value vectors, generated using learnable weight matrices 𝑊^𝑄^, 𝑊^𝐾^ and 𝑊^𝑉^. Here, 𝑄𝐾^𝑇^represents the similarity between patches, with 𝑑_𝑘_as the scaling factor to avoid overly large values, and *Softmax* ensures attention weights sum to 1.

##### 2.2 Multi-Head Attention Capturing Rich Feature Patterns

Multi-head attention enhances feature diversity by processing inputs through multiple attention heads, each capturing distinct patterns. Their outputs are combined as:

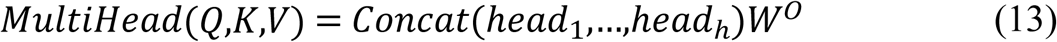

Where each attention head ℎ𝑒𝑎𝑑_𝑖_ is computed as:

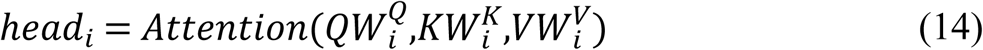

Where 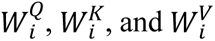 as the weight matrices for the *i*-th head, and 𝑊^𝑂^ as the output projection matrix.

#### C. Multi-View Distillation with BiFusion Blocks

To enhance cross-modal interaction, the BiFusion module integrates multi-scale structural and functional features along with their shared semantic projections, reinforcing structure–function complementarity. BiFusion takes two inputs: fMRI features F from GRU and their shared-space representation ***Z^F^***, and structural features S from CNN with ***Z^S^***.

##### 3.1 Channel-wise Fusion (Functional Modality)

Hadamard product, element-wise addition, and concatenation of ***F_N_*** and its shared representation ***Z_L_***:

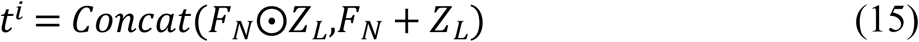

##### 3.2 Channel-wise Fusion (Structural Modality)

Similarly, for ***S_N_*** and ***Z_L_***:

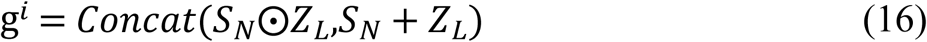

##### 3.3 Attention Modulation (Functional Side)

Global average pooling on 𝑡^𝑖^, followed by a fully connected layer and Sigmoid activation:

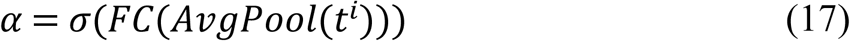

Weighted functional features:

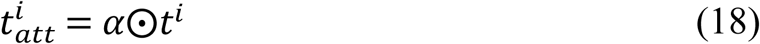

##### 3.4 Attention Modulation (Structural Side)

Apply correlation-based pooling on g^𝑖^, followed by Sigmoid activation:

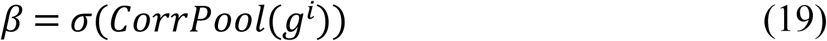

Weighted structural features:

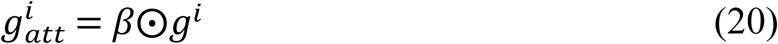

##### 3.5 Fusion and Output Generation

Apply learnable transformations ***W_1_*** and ***W_2_***, followed by convolution and residual connection to obtain the final fused representation:

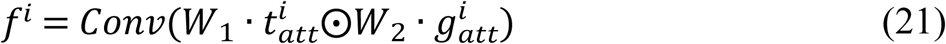

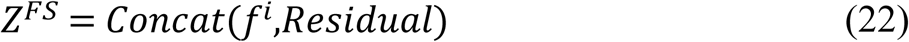

#### D. Progression Prediction Modeling via Multi-Scale Upsampling

To enable accurate disease grading and score prediction, SFNFA-MVD employs a multi-scale upsampling path to progressively restore spatial resolution, with classification and regression outputs generated through prediction heads. The upsampling process is as follows:

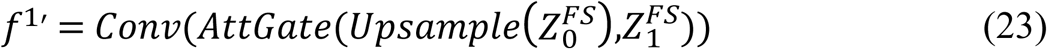

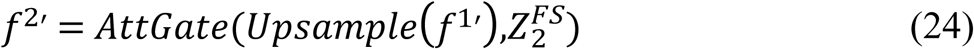

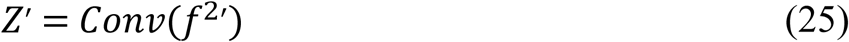

Where *Upsample* increases the resolution of fused features to match the input image size. 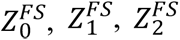 are features processed by the BiFusion module at different resolutions. *AttnGate(*⋅) enhances important feature channels while suppressing irrelevant ones. *Conv(*⋅) extracts finer spatial features, and *Z′* is the final high-resolution output obtained by merging multi-scale features. A convolutional layer then refines the spatial details and reduces the channel count.

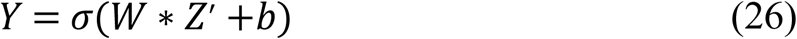

Where *W* is the convolution kernel, ∗ denotes the convolution operation, *b* is the bias term, and 𝜎 is the activation function (e.g., sigmoid), yielding the probabilistic output *Y*, representing the probability of each pixel belonging to a specific class. To further optimize multi-scale features, the prediction head applies a deep supervision mechanism:

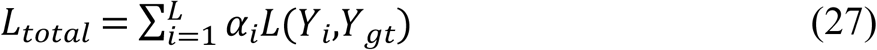

Where 𝑌_𝑖_ is the segmentation output at layer *i*, 𝑌_𝑔𝑡_ is the ground truth, and 𝛼_𝑖_ is the weight for each layer’s loss.

#### E. PD Progression Prediction and Brain Region Localization

##### 4.1 Using L1 Loss for MDS-UPDRS Prediction Error

L1 loss (Mean Absolute Error, MAE) evaluates the difference between predicted and actual MDS-UPDRS scores. It is robust to outliers, unlike L2 loss, and minimizes error amplification. Minimizing L1 loss improves prediction accuracy, better reflecting motor symptom severity. The formula is:

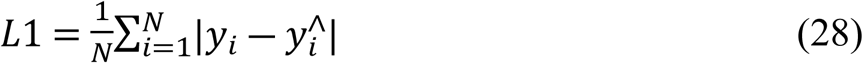

Where 𝑦_𝑖_ is the actual MDS-UPDRS score for sample *i*, 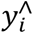 is the predicted score, and *N* is the number of samples.

##### 4.2 Identifying Key Brain Regions with Grad-CAM

To enhance interpretability, SFNFA-MVD uses Grad-CAM to visualize activation heatmaps and identify key brain regions affecting predictions. It calculates gradients of convolutional features for the target class and applies global average pooling for weights. Grad-CAM highlights regions that most influence MDS-UPDRS score predictions by generating a heatmap based on gradient averages from a specific convolutional layer. The principle is as follows:

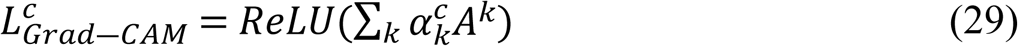

Where 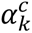 is the gradient weight for motor severity *c* after global average pooling, and 𝐴^𝑘^ is the *k*-th feature map. The ReLU function ensures the heatmap highlights only regions with a positive contribution to the prediction. This approach visualizes brain regions that most influence MDS-UPDRS scores, providing deeper insights into the link between neuroimaging and motor symptoms.

## Results

### A. Performance of SFNFA-MVD Model in Predicting PD Severity

#### 1. Embedding Selection: Optimized Structural and Functional Representations

To determine the optimal unimodal feature encoders for MRI data, we benchmarked multiple architectures for sMRI, fMRI, and DTI across UPDRS-II and UPDRS-III prediction tasks. Performance was evaluated using mean absolute error (MAE) and cumulative score (CS) metrics, reflecting predictive accuracy and clinical reliability, respectively.

For structural embedding, RegNet yielded the most robust representations for both sMRI and DTI encoders. Compared with VGG16-sMRI, RegNet-sMRI achieved a 20.5% reduction in MAE (from 4.532 to 3.602), a 65.3% improvement in CS2 (from 0.436 to 0.721), and a 3.7% improvement in CS3 (from 0.783 to 0.812) for UPDRS-II prediction. A similar pattern was observed for UPDRS-III prediction, where RegNet-sMRI outperformed VGG16-sMRI by 22.0% in MAE (from 4.521 to 3.522), 74.0% in CS2 (from 0.428 to 0.745), and 3.6% in CS3 (from 0.786 to 0.814). Likewise, compared with VGG16-DTI, RegNet-DTI reduced MAE by 19.9% (from 4.612 to 3.693), improved CS2 by 69.6% (from 0.421 to 0.714), and increased CS3 by 3.9% (from 0.772 to 0.802) for UPDRS-II prediction. For UPDRS-III prediction, RegNet-DTI outperformed VGG16-DTI by 22.3% in MAE (from 4.521 to 3.513), 69.9% in CS2 (from 0.432 to 0.734), and 3.2% in CS3 (from 0.784 to 0.809).

For fMRI embedding, GRU outperformed all other encoders in both UPDRS-II and UPDRS-III prediction tasks. Compared with VGG16-fMRI, GRU-fMRI achieved a 26.1% lower MAE (from 4.611 to 3.409), a 62.6% increase in CS2 (from 0.431 to 0.701), and a 0.9% increase in CS3 (from 0.774 to 0.781) for UPDRS-II prediction. Similarly, for UPDRS-III prediction, GRU-fMRI showed a 25.6% reduction in MAE (from 4.451 to 3.312), a 64.3% increase in CS2 (from 0.435 to 0.715), and a 1.0% gain in CS3 (from 0.784 to 0.792).

#### 2. Superior Fusion Strategy: Shared Space with Structural-functional Alignment

Building upon the optimized embeddings, SFNFA-MVD introduces a superior fusion strategy that combines contrastive learning-based shared space with structural-functional aligned integration, which significantly improves cross-modal representation learning for PD progressing modeling. As shown in Table 1, SFNFA-MVD outperformed CLIP by 30.3% in MAE and 15.7% in CS3 for UPDRS-II prediction, and by 28.1% in MAE and 3.0% in CS3 for UPDRS-III. Similarly, compared to TransFuse, SFNFA-MVD reduced MAE by 19.5% (UPDRS-II) and 15.6% (UPDRS-III), while improving CS3 by 3.1% and 4.2%, respectively. These improvements also exceeded those of intermediate fusion strategies, where TransFuse improved upon DeepLabV3+ by up to 24.2% in MAE and 10.6% in CS3 for UPDRS prediction, while CLIP surpassed BLIP-2 by up to 4.9% in CS3.

**Table 1:**
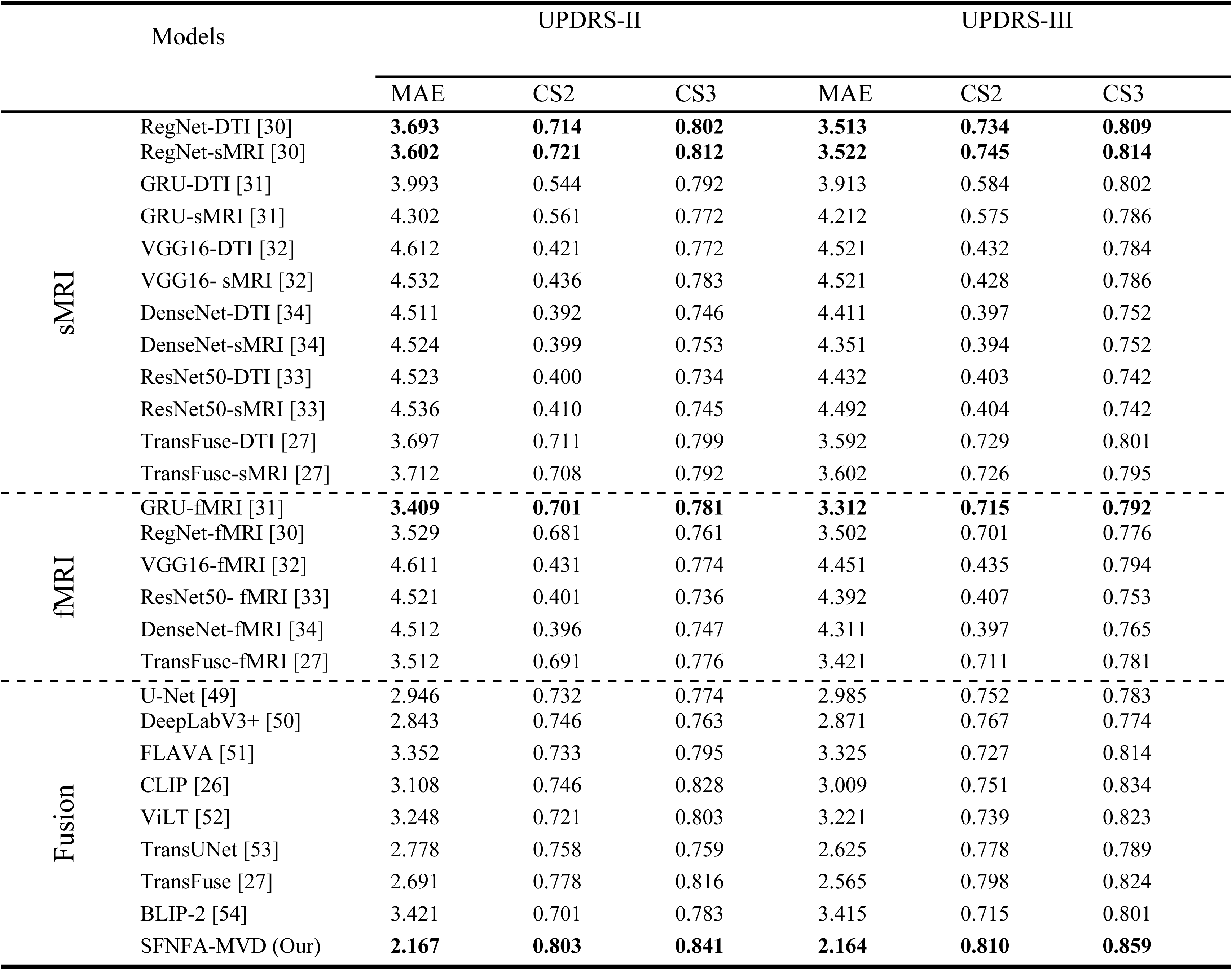
Comparative performance of unimodal and multimodal deep learning models on the PPMI cohort for predicting MDS-UPDRS Part II and Part III scores.

#### 3. Enhanced Prediction Reliability and Multimodal Feature Coherence

To further validate model accuracy and robustness, we examined the correlation between predicted and observed MDS-UPDRS scores across different architectures (Figure 2). SFNFA-MVD achieved the highest Pearson correlation coefficients among all models, with r = 0.613 for UPDRS-II prediction and r = 0.695 for UPDRS-III prediction (p < 0.001), outperforming unimodal baselines including both GRU-fMRI and RegNet-sMRI. These results indicate the enhanced predictive reliability of SFNFA-MVD, underscoring its capacity to model latent clinical trajectories with both sensitivity and stability.

**Figure 2.**
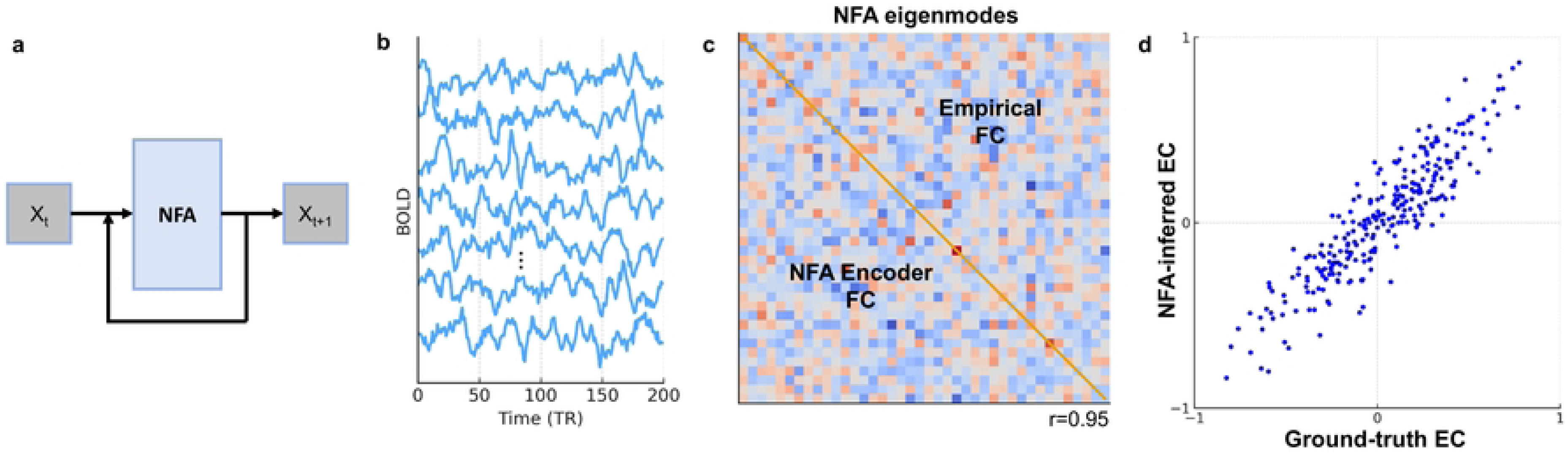
Comparison of model performance in predicting PD motor symptoms as measured by UPDRS-II and UPDRS-III. The x-axis represents the observed UPDRS scores, and the y-axis represents the predicted values. Each dot denotes one subject, with the solid line indicating the linear regression fit and the shaded area showing the 95% confidence interval. The SFNFA-MVD model achieved the highest correlations for both UPDRS-II (r = 0.613, p < 0.001) and UPDRS-III (r = 0.695, p < 0.001), outperforming the unimodal baselines RegNet-sMRI (r = 0.507/0.599) and GRU-fMRI (r = 0.542/0.662).

### B. Discovery of Functional Network Subtypes in PD Progression

#### 1. Functional Network Patterns Across PD Progression Rates

To investigate whether SFNFA-MVD captures neurobiologically meaningful distinctions among PD progression trajectories, we conducted unsupervised clustering on the model-derived multimodal embeddings using t-distributed stochastic neighbor embedding (t-SNE) and applied established progression subtyping criteria[11]. This analysis revealed three distinct clusters corresponding to slow-progressing (SP, green), moderate-progressing (MP, blue), and fast-progressing (FP, red) subtypes (Figure 3). Each cluster was associated with a distinct functional connectivity profile anchored in large-scale cortical networks. The SP group exhibited connectivity patterns predominantly centered on the sensorimotor network (SMN); the MP group exhibited network features aligned with the dorsal attention network (DAN); and the FP group showed a dominant pattern involving the salience network (SN). The identification of these subtypes highlights SFNFA-MVD’s capability to uncover latent brain network signatures that differentiate PD trajectories.

**Table 2:**
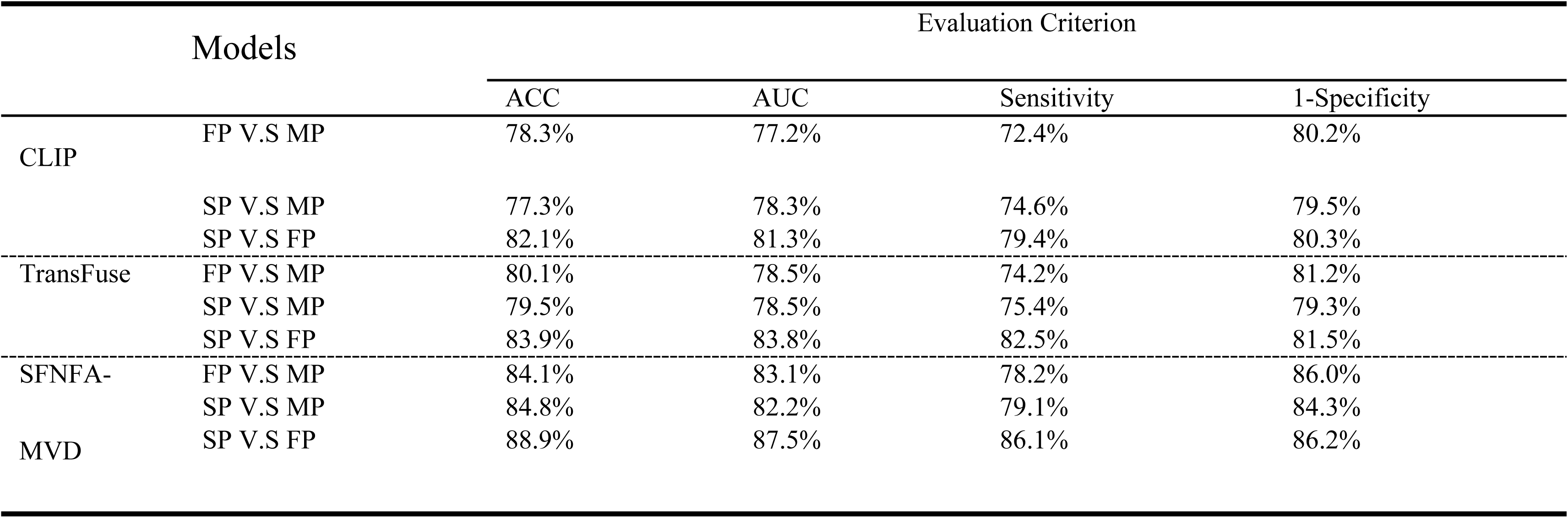
Performance of the SFNFA-MVD framework in classifying PD progression subtypes based on multimodal MRI features. FP: fast progressing; MP: moderate progressing; SP: slow progressing.

**Figure 3.**
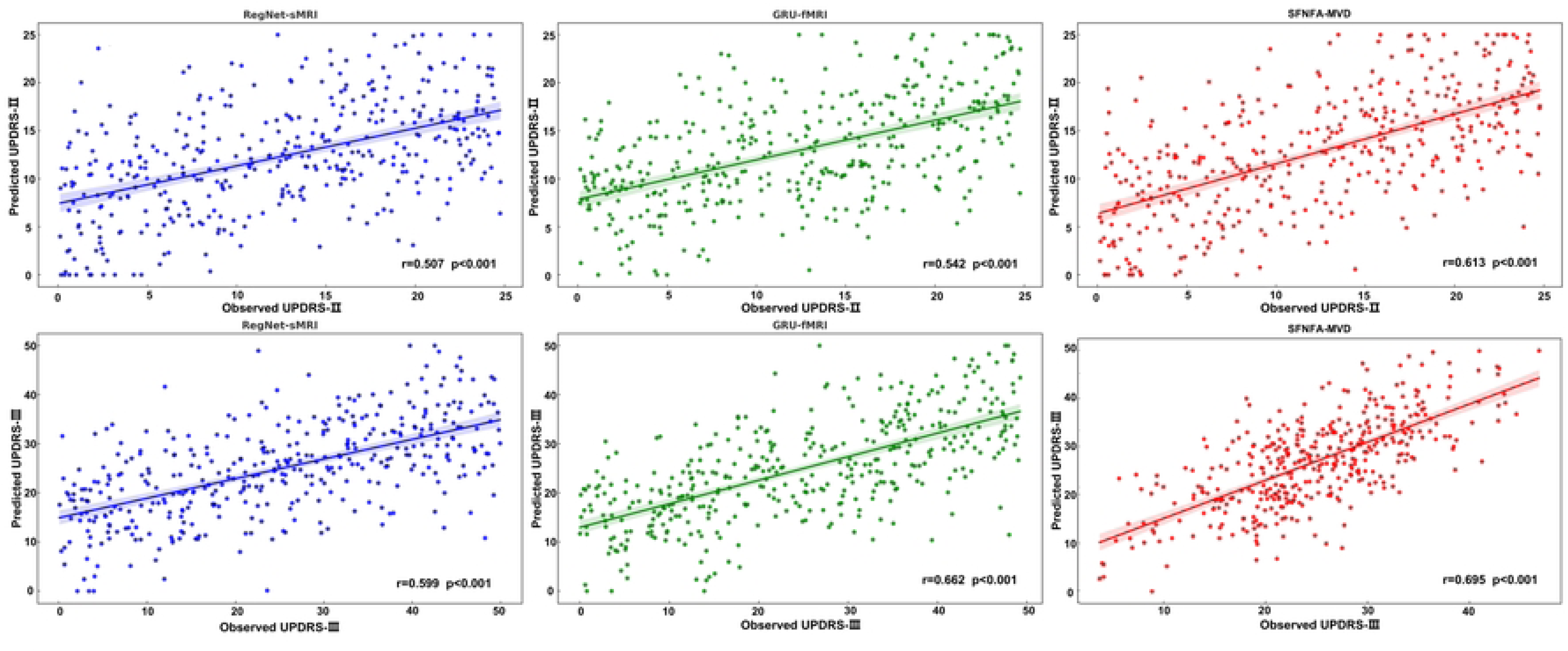
Functional network density and cortical distribution across PD progression subtypes. *(A)* Cortical surface maps illustrate the dominant network clusters identified in the slow-progressing (SP), moderate-progressing (MP), and fast-progressing (FP) subgroups. Color intensity indicates the relative strength of network connectivity, highlighting spatially distinct patterns across progression phenotypes: enhanced sensorimotor (SMN) engagement in SP, strengthened dorsal attention network (DAN) in MP, and prominent salience network (SN) activation in FP groups. *(B)* Violin–box plots depict quantitative differences in network density among subgroups. Each dot represents an individual participant, and the boxplots summarize the distribution of SMN, DAN, and SN connectivity densities. Statistical comparisons show significant group differences (p < 0.001) across all three networks, revealing a gradual shift from sensorimotor to salience-dominant organization as disease progression accelerates.

**Table 3:**
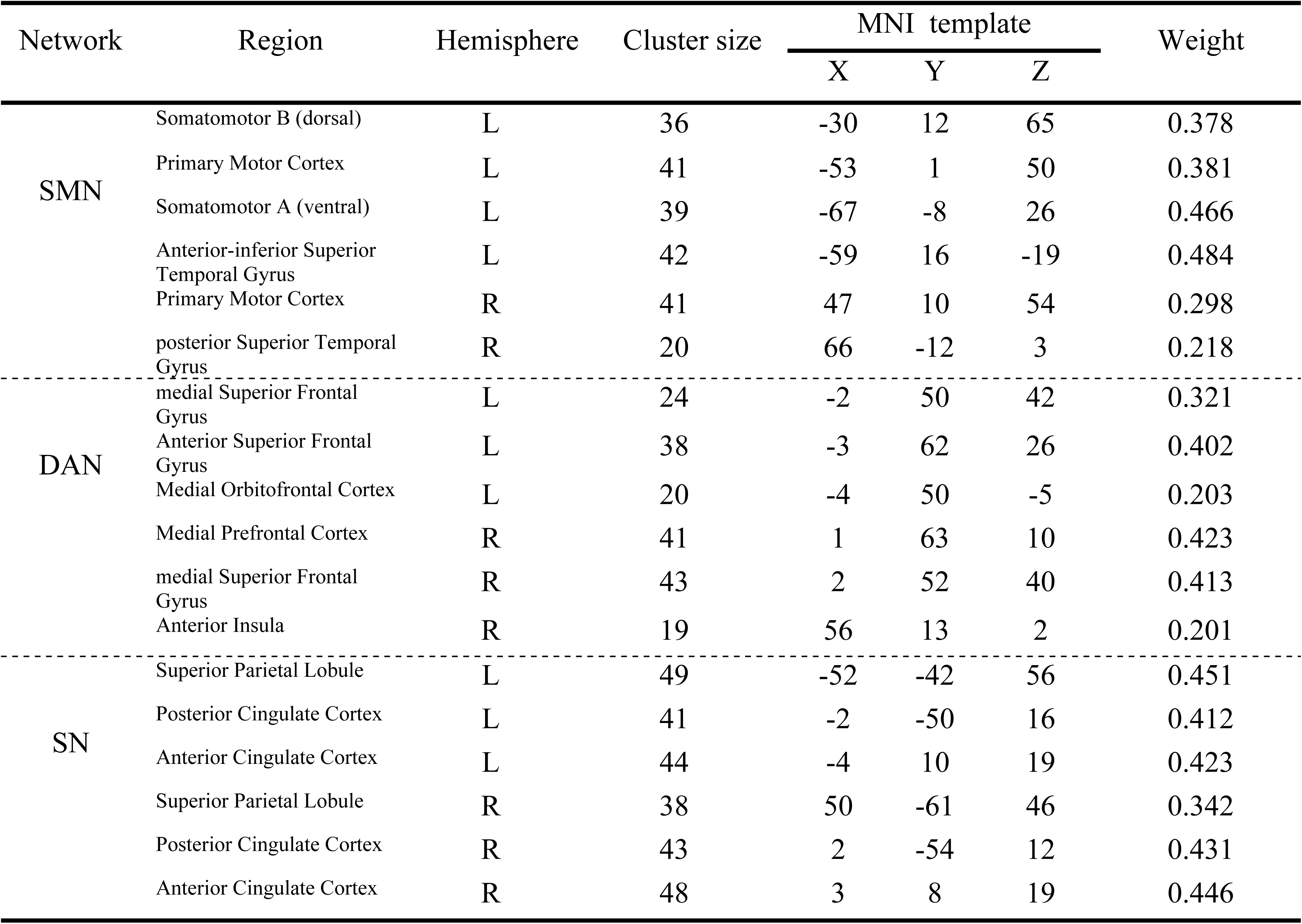
Peak coordinates and corresponding cortical regions of the SP, MP, and FP clusters. FP: fast progressing; MP: moderate progressing; SP: slow progressing.

#### 2. Functional Correlates of PD Progression Prediction

To elucidate the contribution of specific brain subnetworks to longitudinal PD progression, we assessed the correlations between functional network density and longitudinal changes in motor symptom severity (MDS-UPDRS Part II and Part III scores). As shown in Figure 4 and Figure 5, distinct functional networks were associated with different progression subtypes. In Figure 4 (UPDRS-II), the SMN exhibited the strongest correlation in the SP group (r = 0.54), the DAN in the MP group (r = 0.55), and the SN in the FP group (r = 0.56). Figure 5 (UPDRS-III) revealed a similar pattern, with SMN showing the highest correlation in the SP group (r = 0.56), DAN in the MP group (r = 0.57), and SN in the FP group (r = 0.56).

**Figure 4.**
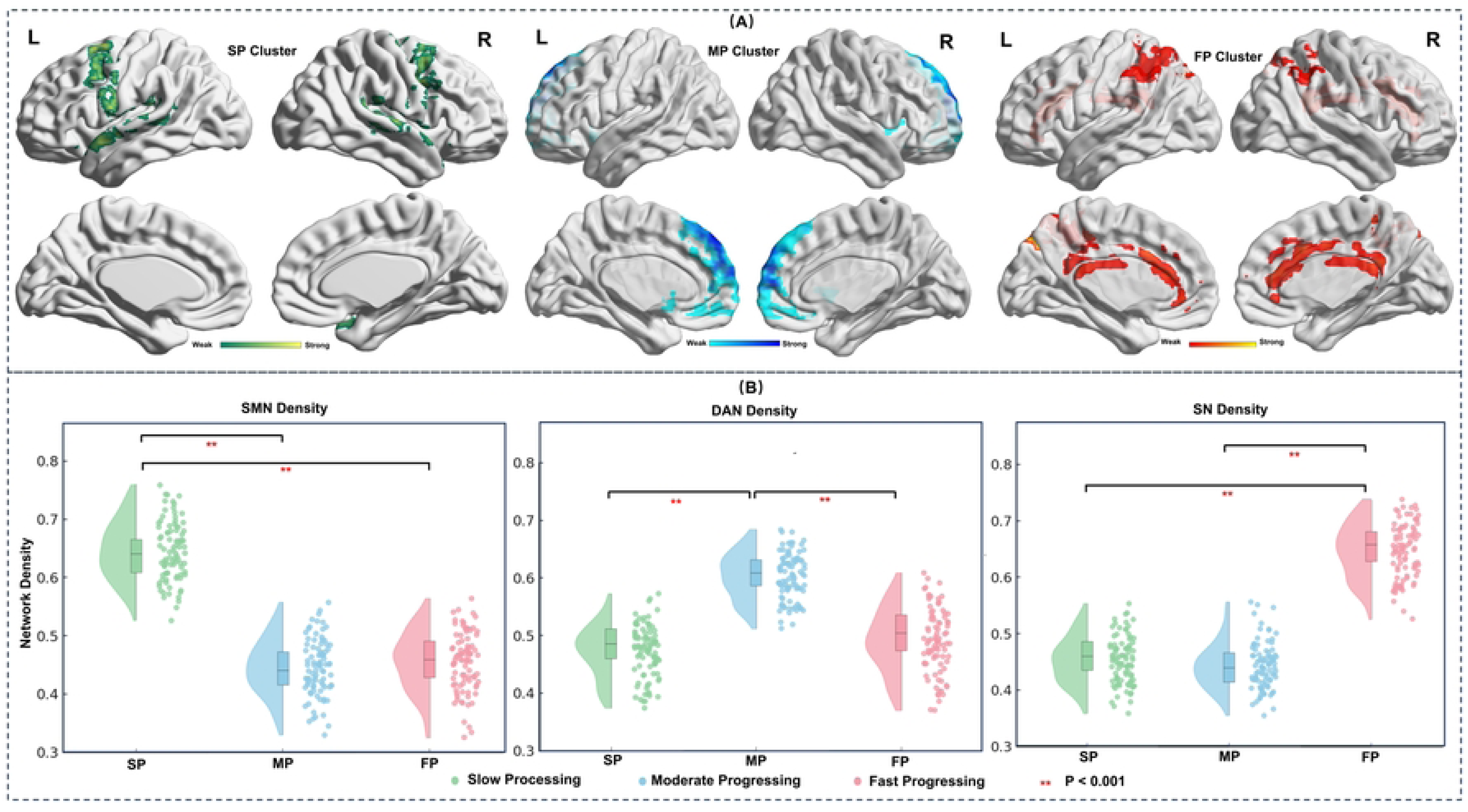
Correlations between functional network density and UPDRS-II progression rate across PD subtypes. Scatter plots illustrate the associations between network density (y-axis) and UPDRS-II progression rate (x-axis) over two follow-up periods (0-3 years, top row; 0-5 years, bottom row) for the three PD progression subgroups: slow-progressing (green, SP), moderate-progressing (blue, MP), and fast-progressing (red, FP). Each column corresponds to a major functional network: sensorimotor network (SMN), dorsal attention network (DAN), and salience network (SN). Across all subgroups and time windows, significant positive correlations (*ps* < 0.001) were observed, with the strongest associations found within the predominant networks of each subtype, including SMN for SP, DAN for MP, and SN for FP.

**Figure 5.**
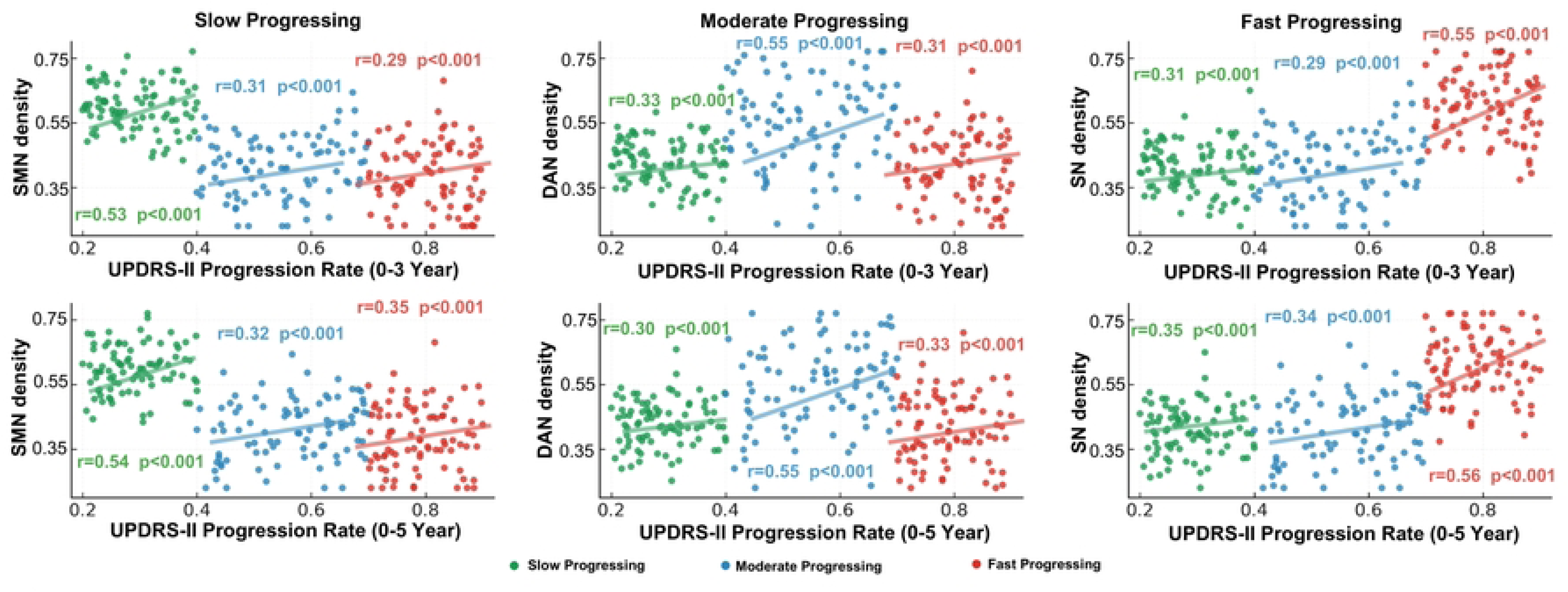
Correlations between functional network density and UPDRS-III progression rate across PD subtypes. Scatter plots show the relationships between functional network density (y-axis) and UPDRS-III progression rate (x-axis) over two follow-up periods (0-3 years, top row; 0-5 years, bottom row) for three Parkinson’s disease subgroups: slow-progressing (green, SP), moderate-progressing (blue, MP), and fast-progressing (red, FP). Each column corresponds to a major functional network: sensorimotor network (SMN), dorsal attention network (DAN), and salience network (SN). Across all subgroups and time windows, significant positive correlations (*ps* < 0.001) were observed, with the strongest associations found within the predominant networks of each subtype, including SMN for SP, DAN for MP, and SN for FP.

### C. Transfer Learning Enables Prediction of Long-Term PD Progression

To evaluate the generalizability of SFNFA-MVD for long-term disease monitoring, we applied a transfer learning strategy that adapts knowledge from short-term (0-3 year) prediction to longer-term (0-5 year) progression. As shown in Figure 4 and Figure 5, functional network density remained a stable and robust predictor in long-term progression estimation. For the FP group, the correlation between the SN and UPDRS-II increased from r = 0.55 to 0.56, representing an improvement of approximately 1.8%. For the MP group, the DAN showed an increase in correlation with UPDRS-III from r = 0.55 to 0.57, corresponding to an improvement of about 3.6%. For the SP group, the SMN increased from r = 0.53 to 0.54 (+1.9%) for UPDRS-II and from r = 0.55 to 0.56 (+1.8%) for UPDRS-III.

## Discussion

The present study developed and validated SFNFA-MVD, a novel deep learning framework that integrates multimodal MRI for individualized prediction of PD progression. By combining localized feature encoding across structural and functional modalities with a transformer-based architecture that captures long-range temporal and spatial dependencies, SFNFA-MVD achieved state-of-the-art accuracy in predicting longitudinal UPDRS-II and UPDRS-III scores, outperforming all benchmark unimodal and multimodal models. Moreover, SFNFA-MVD revealed three progression subtypes, including SP, MP, and FP subtypes, characterized by distinct network patterns along a hierarchical gradient extending from SMN to DAN and SN. Importantly, functional network density within the SMN, DAN, and SN showed significant correlations with longitudinal motor symptoms. Together, these findings establish SFNFA-MVD as a robust and clinical interpretable framework for modeling the heterogeneity of PD progression, highlighting the importance of multimodal integration for precision modeling of neurodegenerative disease.

SFNFA-MVD achieved state-of-the-art accuracy in predicting PD motor symptoms, reducing MAE by up to 30.4% and improving CS3 by up to 12.7% compared to unimodal models (e.g. RegNet-sMRI, RegNet-DTI, and GRU-fMRI). It also outperformed advanced multimodal fusion architectures including CLIP and TransFuse, achieving the highest prediction consistency across longitudinal time points, reducing MAE by up to 30% and improving CS3 by up to 16% compared with CLIP, and further lowering MAE by ∼20% and raising CS3 by 3–4% relative to TransFuse. These improvements were further supported by strong prediction-observation correlations for both UPDRS-II (r = 0.489, p < 0.001) and UPDRS-III (r = 0.494, p < 0.001). In contrast, contrastive variational autoencoders using sMRI achieved moderate classification accuracy (AUCs=0.66–0.75) for short-term stage transitions [36]. Similarly, a graph-based multimodal approach that combined sMRI with clinical and genetic data reported AUCs of 0.748 for H-Y stage and 0.714 for UPDRS-III prediction at 12 and 36 months [37]. Our study advances beyond these findings by providing evidence that integrating structural and functional neuroimaging modalities can substantially enhance accuracy and robustness in predicting PD motor symptoms.

Building upon its superior performance in predicting motor symptoms, SFNFA-MVD further identified three subtypes of PD progression through unsupervised clustering of multimodal MRI fusion. Each subtype was characterized by a distinct functional network, reflecting a hierarchical gradient of degeneration from the SMN in the SP subtype to the DAN in the MP subtype and the SN in the FP subtype. More importantly, the validity of these subtypes was confirmed by significant correlations between functional network density and longitudinal motor symptom. Specifically, functional connectivity density within the SMN, DAN, and SN showed significant correlations with longitudinal changes in motor symptoms for the SP (UPDRS-II: r = 0.53; UPDRS-III; r = 0.56), MP (UPDRS-II: r = 0.54; UPDRS-III: r=-0.57), and FP (UPDRS-II: r = 0.52; UPDRS-III: r = -0.56) subtypes, respectively. This network-specific correspondence between functional connectivity and motor progression provides direct support for the biological validity of the SFNFA-MVD-derived progression subtypes.

Previous studies have characterized PD heterogeneity primarily through clinical time-course modeling or probabilistic state-transition analyses [10, 11, 18], which identified SP and FP subtypes but lacked imaging correlates to support these distinctions. Similarly, biomarker- and symptom-based clustering models improved clinical differentiation by integrating genetic or fluid measures [38, 39] but offered limited insight into the network-level organization. More recently, fusion of MRI and DAT-SPECT achieved classification accuracies around 78-79% in distinguishing progression groups [40], but they did not characterize the neural substrates underlying motor decline. In contrast, SFNFA-MVD identifies progression subtypes through multimodal MRI fusion and validates them through significant correlations between network integrity and behavioral deterioration, thereby providing a neuroimaging framework for characterizing the clinical heterogeneity and trajectory diversity of PD progression.

The SMN-DAN-SN hierarchical gradient identified by SFNFA-MVD represents a biologically plausible trajectory of PD progression. This organization consists of Braak’s staging of α-synuclein propagation and contemporary network-degeneration models, in which pathology spreads from subcortical motor circuits to transmodal cortical hubs via structurally and functionally connected pathways [41, 42]. Specifically, the SP subtype showed early degeneration in the M1 and PMC within the SMN, reflecting localized motor dysfunction with relative preservation of higher-order regions [43]. The MP subtype exhibited disruption of the PMC and parietal components of the DAN, suggesting impaired visuomotor integration and attentional control that may initially serve compensatory roles but deteriorate with progressive frontoparietal disconnection [44]. In contrast, the FP subtype showed prominent alterations in the STG and SN. These regions are integral to network switching, cognitive control, and emotional regulation and their damage has been associated with rapid cognitive and affective decline in PD [45]. This hierarchical gradient also aligns with connectomic vulnerability models [46], which posit that transmodal hubs (e.g. the DAN and SN) are more susceptible to synuclein accumulation and metabolic stress due to their high centrality and integrative demands. Consistent with these models, neuroimaging studies have shown that localized SMN disruption precedes widespread fronto-insular and limbic dysfunction as disease severity increases [43, 47]. Together, the SFNFA-MVD-derived gradient reveals a mechanistic progression from localized motor dysfunction to widespread limbic network disintegration, offering an interpretable framework for understanding the neural basis of PD heterogeneity.

The superior prediction and subtyping performance of SFNFA-MVD arises from its architectural advantages over existing unimodal and multimodal approaches. Previous PD models typically rely on single-modality features or combine modalities through simple feature concatenation that fail to capture the interdependencies between structural integrity and functional reorganization [10, 48]. SFNFA-MVD addresses this gap through two complementary modules: the CLIP and the TransFuse. The CLIP module performs fine-grained alignment of regional features between structural and functional representations, projecting regional morphology, connectivity strength, and temporal dynamics into a shared latent space. Building upon these aligned embeddings, the TransFuse module employs a transformer-based attention mechanism to model long-range dependencies across brain regions and modalities, enabling hierarchical integration of both localized and distributed neural features. Together, these modules allow SFNFA-MVD to model the multiscale architecture of PD from localized motor disruptions to widespread network disintegration, providing precise and interpretable representations that support both individualized prediction and progression subtyping.

Several limitations should be considered when interpreting these findings. First, this study was conducted within a single cohort with limited ethnic and clinical diversity, which may restrict the generalizability of SFNFA-MVD across broader PD populations. Future validation using independent, multi-site datasets will be essential to confirm robustness and reproducibility. Second, this study focused exclusive on motor symptoms (MDS-UPDRS II and III), and future work should extend to non-motor domains to provide a more comprehensive understanding of PD progression. Finally, although SFNFA-MVD identified clinically plausible progression subtypes, the relationships between network-level alterations and motor decline remain correlational. Longitudinal multimodal studies with causal modeling or intervention designs are necessary to establish mechanistic pathways underlying subtype-specific progression.

## Conclusion

In summary, SFNFA-MVD provides a multimodal deep learning framework that enables individualized modeling of PD progression. The model not only achieves state-of-the-art accuracy in predicting longitudinal motor symptom but also identify three progression subtypes characterized by network-specific patters along a sensorimotor-attentional-salience gradient. By integrating structural and functional MRI within a unified representation, SFNFA-MVD establishes a clinically interpretable framework for capturing the heterogeneous trajectories of PD progression.

## Data Availability

The data that support the findings of this study are publicly available from the PPMI dataset.

## Data and code availability

The multimodal MRI data used in this study were obtained from the Parkinson’s Progression Markers Initiative (PPMI). Access to the dataset requires data-use agreement and institutional approval through https://www.ppmi-info.org. Derived data supporting the findings of this study and pretrained model weights are available from the lead contact upon reasonable request for research collaboration. The complete source code for SFNFA-MVD and related scripts is publicly available on GitHub: https://github.com/zhaoshuhzi/PD_PPMI_S-F_Fusion.

## Acknowledgements

We thank all members of the Nan Yan and Hanjun Liu laboratories for valuable discussions and technical assistance. This work was supported by the National Natural Science Foundation of China (U23B2018, 62271477, 82371471, 82472600, 82172528), the Shenzhen Science and Technology Program (JCYJ20220818101217037, KQTD20200820113106007, JCYJ20240813151223031), and the Shenzhen Medical Research Fund (C2401001).

## Author contributions

Conceptualization, S.Z. and N.Y.; methodology, S.Z., Y.Z., and N.Y.; software, S.Z. and Y.Z.; validation, S.Z. and M.L.N.; formal analysis, S.Z. and H.L.; investigation, S.Z., Y.Z., and L.W.; resources, N.Y. and L.W.; data curation, S.Z. and Y.Z.; writing – original draft, S.Z.; writing – review & editing, S.Z., M.L.N., H.L., L.W., and N.Y.; supervision, N.Y.; project administration and funding acquisition, N.Y.

## Competing interests

The authors declare no competing interests.

---

**Figure 6.**
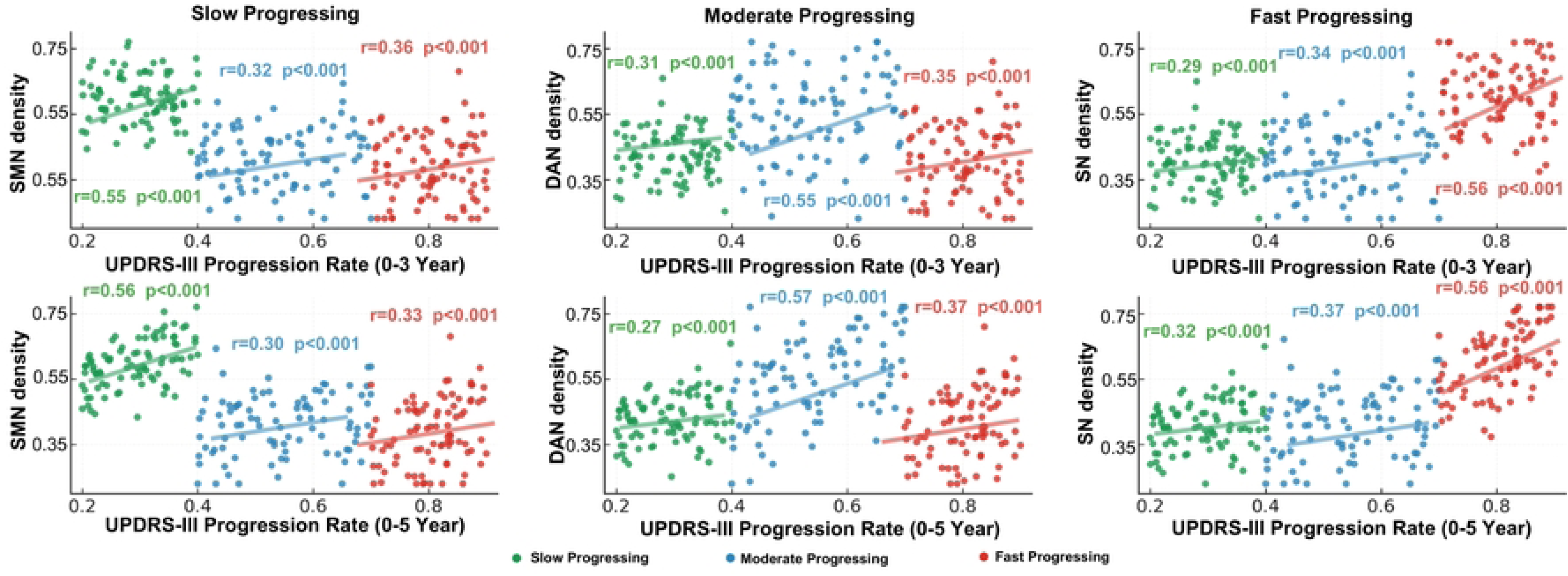

## Notes

### Competing Interest Statement

The authors have declared no competing interest.

### Funding Statement

Yes

### Author Declarations

The First Affiliated Hospital, Sun Yat-sen University Medical Ethics Committee

